# Noninvasive prenatal exome sequencing inefficient for detecting single-gene disorders – problems and possible solutions

**DOI:** 10.1101/2020.08.04.20168278

**Authors:** Dayne L Filer, Piotr A Mieczkowski, Alicia Brandt, Kelly L Gilmore, Bradford C Powell, Jonathan S Berg, Kirk C Wilhelmsen, Neeta L Vora

**Affiliations:** Department of Genetics, School of Medicine, UNC Chapel Hill, Chapel Hill, NC; Renaissance Computing Institute, Chapel Hill, NC; Department of Obstetrics & Gynecology, School of Medicine, UNC Chapel Hill, Chapel Hill, NC; Department of Neurology, School of Medicine, UNC Chapel Hill, Chapel Hill, NC

## Abstract

**What’s already known about this topic?:**

- Sequencing-based noninvasive testing can detect large copy number abnormalities and some autosomal dominant single-gene disorders
- Exome sequencing (ES) on fetal samples provides 20% diagnostic yield for structural abnormalities after normal karyotype & microarray

**What does this study add?:**

- ES on cell-free DNA in three gravid patients with suspected genetic disease in the fetus
- We demonstrate broad sequencing approaches are limited by sampling and technical difficulties, concluding broad sequencing is currently inappropriate for noninvasive testing

## Letter

The beneficial health outcomes from newborn screening programs (NBS) are indisputable. We envision is future NBS will begin with prenatal genetic testing to enable care in the immediate newborn period, and open up new possibilities for *in utero* and genetic therapies. During pregnancy placental DNA is released into maternal circulation, enabling noninvasive interrogation of fetal genetics (noninvasive pre-natal testing, NIPT). NIPT has a well-established clinical utility in screening for common chromosomal abnormalities such as Down syndrome with high sensitivity and specificity.^1^ More recently, efforts have demonstrated sequencing-based testing for *de novo* pathogenic variants in a list of 30 genes associated with dominant Mendelian disorders^2^ and PCR-based testing for a small number of recessive Mendelian disorders.^3^ To date, no one has reported reliable fetal genotyping purely from maternal cell-free DNA using a sequencing-based approach.

To begin NBS with prenatal genetic testing, we believe we first need a reliable noninvasive test only requiring a maternal sample. Others could reasonably argue the availability of carrier screening, and the immeasurably small risk of invasive testing,^4^ removes the need for the noninvasive test. Such an argument, however, dismisses (1) the ethical and practical issues surrounding the necessity of involving the biological father, (2) the fact that many genetic disorders arise due to *de novo* mutations, and (3) the understandable fear and apprehension around invasive testing (especially for rare conditions). Additionally, we believe the prenatal diagnosis community should focus work on sequencing-based (as opposed to PCR-based) approaches. Sequencing-based approaches generalize across disorders more easily than PCR-based approaches, multiplex to a degree not feasible using PCR, and will only continue to decrease in cost.

Previously, Kitzman et al. performed whole-genome fetal sequencing from maternal plasma by combining whole genome data from the father, mother, and maternal plasma,^5^ but illustrate the cost-infeasibility in their subsequent review article and suggest more targeted approaches such as exome sequencing (ES).^6^ As an exploratory exercise, we performed ES on cell-free DNA (cfES) from three pregnant women with singleton fetuses.

Briefly, we collected cell-free DNA from maternal plasma, prepared sequencing libraries for the Illumina platform, and performed exome capture using the IDT xGen Exome Research Panel v1.0 (Cases 1 & 2) or Agilent SureSelect Human All Exon v7 (Case 3). All participants were consented and enrolled at UNC Hospitals by certified genetic counselors with approval from the UNC Institutional Review Board (IRB Number: 18-2618); we do not include any identifying information in this manuscript. We processed the data using a novel analytic pipeline developed in Snakemake using Anaconda environments for reproducibility. Sequencing reads were aligned to hg38 (excluding alternate contigs) using BWA-MEM, then base quality scores were re-calibrated using GATK4. We only retained non-duplicate, properly-paired reads with unambiguous mapping and mapping quality >30 for each read. We called variants using the bcftools software requiring basepair quality scores >20, 5 alternate allele-supporting fragments, and 80 total fragments. Analyses were restricted to the regions overlapping between the IDT and Agilent capture platforms. Using the identified single-nucleotide variants, we applied a novel empirical Bayesian procedure to estimate the fetal fraction (FF; the proportion of placental/fetal to maternal sequencing reads). We then estimated fetal and maternal genotypes using a maximal likelihood model incorporating the FF estimate and observed proportion of minor allele (alternate) reads (PMAR). Full analytic pipeline available upon request.

Table 1 lists the known genetic diagnoses for the three cases presented. Genetic counselors recruited the three participants with investigators and cfES analysis blinded to the eventual genetic diagnoses. In Cases 1 & 2, specific gene sequencing based on family history and sonographic findings, respectively, provided genetic diagnoses. To date, Case 3 does not have a specific genetic diagnosis despite whole-genome sequencing of the newborn and ES on the trio. Afterwards, we learned the mother in Case 1 carries a deletion of exon 1 in the gene most-often responsible for Menke’s syndrome (ATP7A). Neither exome capture platform targets ATP7A exon 1; therefore, cfES could not have identified the diagnosis for Case 1 with the platform used. In Case 2, we identified the causal variant using cfES. In this case, we correctly genotyped the fetus, but lacked the power to make the genotyping call with any level of confidence acceptable for clinical use (fig. 1B, note the widely-overlapping distributions at the causal variant). We did not identify any known pathogenic variants in the sequencing of Case 3, and despite performing whole-genome sequencing on the newborn, we still do not have a genetic diagnosis for the family.

**Table 1:** Case summaries. GA: gestational age at the time of blood draw for cfES. FF: estimated fetal fraction. Depth: median depth used to estimate genotypes (does not include duplicated/filtered reads). %Dup: percentage of total mapped read pairs discarded as PCR and/or optical duplicates. %Filt: percentage of total mapped read pairs discarded for improper pairing and/or mapping quality.

|  | GA | Clinical findings | Genetic diagnosis | FF | Depth | %Dup | %Filt |
| --- | --- | --- | --- | --- | --- | --- | --- |
| 1 | 32w2d | 5 prior pregnancies affected with X-linked recessive Menke's syndrome | Menke's syndrome; del. ATP7A exon 1 | 0.117 | 241 | 42.8 | 21.96 |
| 2 | 24w5d | Fetal sonogram at 21w5d showed femoral bowing with shortened length (<3% for GA) bilaterally | Osteogenesis imperfecta type VIII; P3H1 c.1120G>T (rs140468248) | 0.122 | 152 | 33.32 | 22.09 |
| 3 | 34w0d | Fetal sonogram at 19w0d showed bilateral club foot with bilateral upper limb arthrogryposis | None, to date, despite exome and genome sequencing of newborn | 0.169 | 330 | 53.67 | 32.65 |

**Figure 1:**
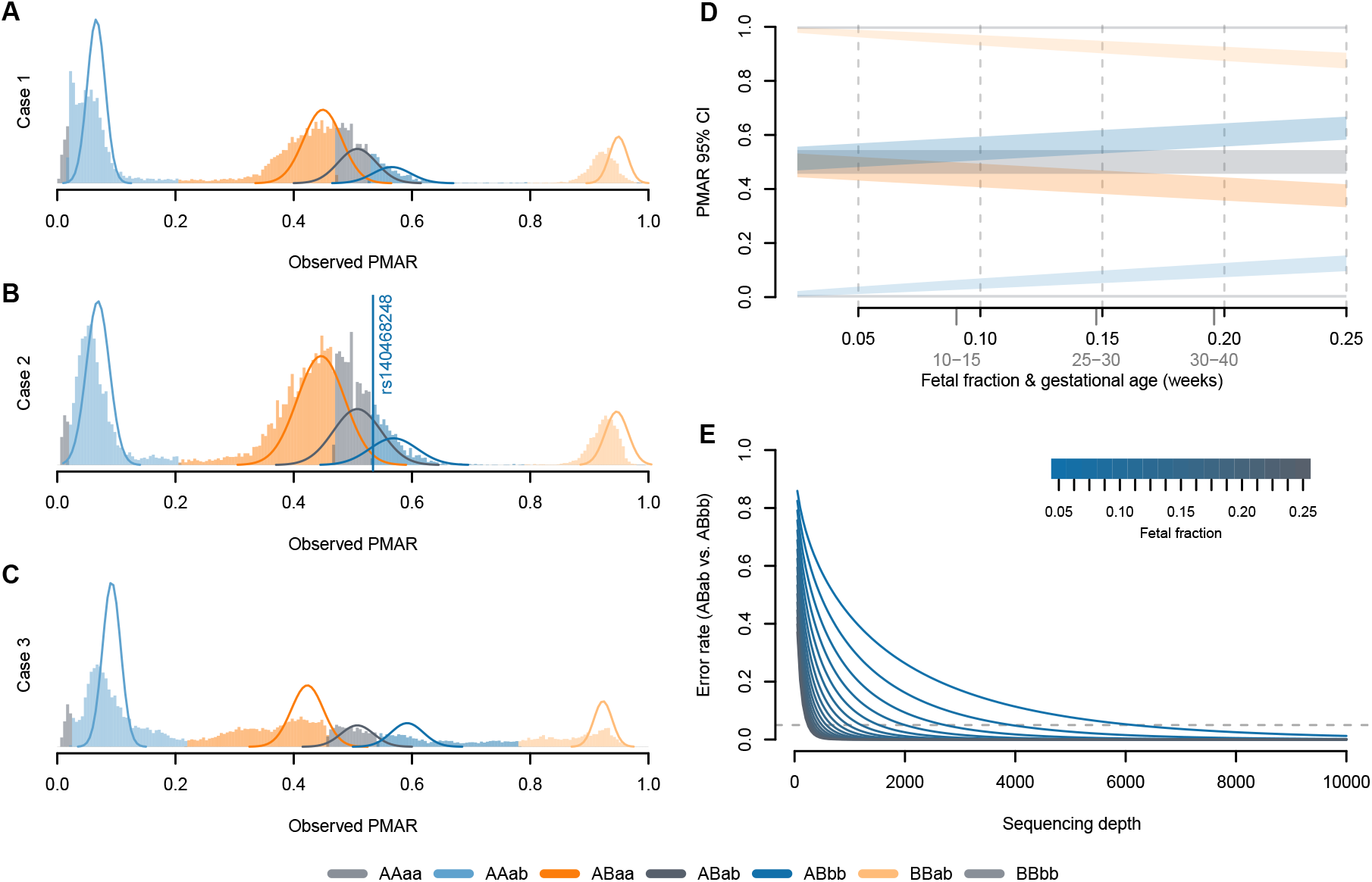
[A-C] Distribution of observed PMAR values for the three cases across the possible maternal-fetal genotype pairs. Uppercase letters give the estimated maternal genotype, lowercase letters give the estimated fetal genotype; ‘A/a’ indicates the reference allele, ‘B/b’ indicates the alternate allele. Solid lines show the normal approximation for the theoretical distribution of binomial probabilities, given the frequency of the estimated genotypes. The vertical line in [B] shows the observed PMAR for the known pathogenic variant, rs140468248. [D] 95% confidence intervals on the binomial proportions for possible maternal-fetal genotype pairs across increasing fetal fractions; represents a sequencing depth of 500x. Average fetal fractions by gestational age (in weeks) given in light gray.^7^ [E] Expected misclassification rate (Weitzman overlapping coefficient; i.e. the area of overlapping distributions in [D]) considering ABab versus ABbb as a function of sequencing depth and fetal fraction. The dashed horizontal line shows 5% error. The theoretical error rates for ABab vs ABaa are symmetric and equal; however, the frequency of errors will depend on the population frequency of the reference versus alternate allele.

Without the ability to reliably exclude maternal DNA fragments, noninvasive sequencing-based methods to genotype the fetus either require additional sequencing of parental samples or distinguishing genotypes by the proportion of minor allele reads (PMAR). Here, we make no attempt to utilize parental genetic information and demonstrate the difficulty of inferring the genotypes directly from the PMAR. We model the PMAR as a binomial proportion; given the fetal fraction, one can prove the true PMAR defines the maternal-fetal genotype combination.

For illustration, consider watching two people randomly place balls into an urn. We know each person either has all white balls, all black balls, or equal numbers of white and black balls; we also know the number, but not the color of balls each person places. We count 60 black balls and 40 white balls in the urn. Given Person A placed 80 balls, the maximum likelihood estimate suggests Person A had equal white and black balls (0.5 × 80 = 40) and Person B had all black balls (1.0 × 20 = 20).

The theoretical bounds of the binomial distribution, therefore, confine our ability to discriminate maternal-fetal genotypes. Using the normal approximation for the binomial variance (valid when the number of observations (sequencing depth), N, times the binomial proportion (PMAR), p is greater than 10), we can clearly explain the poor results we observed (fig. 1D-E). At sequencing depths up to 500x, the 95% confidence intervals on PMAR distributions still overlap for fetal fractions up to roughly 0.17 (fig. 1D). When we calculate the degree of distribution overlap (a proxy for classification error rate), we see required sequencing depths in excess of 8,000x for low fetal fraction samples.

The sequencing herein likely suffers from three problems: (1) inadequate sequencing depth; (2) biased PMAR values from the removal of duplicate reads; (3) errors in sequencing and/or PCR. We have already illustrated the inadequate depth, but emphasize that the theoretical results we present speak to the final depths (not the raw sequencing depth). In our three cases, we excluded over half the reads taken off the sequencer due to sequencing quality thresholds (table 1). We observe the evidence of problems (2) and (3) by observing the high proportion of both duplicate reads and PMAR values outside the theoretic distributions. Additionally, for Case 3 only, we can assess the accuracy of the genotype estimates. In Case 3, we have ES from newborn cord-blood; if we examine variants from both the cfES and ES of newborn cord-blood, we observe a 50.9% genotyping accuracy (data not shown).

Typical sequencing workflows start with randomly fragmenting DNA molecules to build sequencing libraries. Standard bioinformatic practices suggest we remove read-pairs with identical endpoints, because the duplicate read-pairs more likely represent PCR amplification of a single molecule than two molecules with the same fragmentation. Cell-free DNA molecules are shorter than nuclear DNA, not requiring manual fragmentation, and have a non-random distribution of endpoints. Therefore, compared to standard sequencing libraries, the likelihood of observing true duplicates in cell-free libraries increases and we cannot necessarily assume duplicates represent PCR amplification. However, for this work we have no way of differentiating reads representing true duplicate molecules versus PCR duplicates and thus excluded duplicate reads from our analysis.

To solve the above issues, we are currently developing and testing a more targeted approach with sequencing depths in excess of 10,000x and unique molecular identifiers to estimate accurately sequencing errors and differentiate true versus artifactual duplicate reads. Given the depth requirements for estimating fetal genotypes by the PMAR, and the challenge of variants of uncertain clinical significance, we advocate against broad sequencing modalities on noninvasive samples. Despite the challenges ranged by this letter, we have good reason to believe we can assess hundreds to thousands of basepairs, rather than the tens of millions targeted in ES, economically and reliably. In doing so, we hope to foster population-level screening for Mendelian disorders during the prenatal period and, ultimately, unlock new avenues in the treatment of these disorders.

## Data Availability

Full analytic pipeline available upon request.

## Acknowledgements

We thank Dr. James Evans for providing review and feedback of this manuscript. We especially thank the authors of the software packages that we did not have room in this brief letter to cite directly. Neeta Vora and this work was supported by NICHD (K23HD088742). Dayne Filer was supported by NICHD (F30HD101228) and by NIGMS (5T32GM067553).

## Word count

~~~
File: ResearchLetter.tex
Encoding: utf8
Words in text: 1577
Words in headers: 18
Words outside text (captions, etc.): 235
Number of headers: 5
Number of floats/tables/figures: 2
Number of math inlines: 5
Number of math displayed: 0
Subcounts:
  text+headers+captions (#headers/#floats/#inlines/#displayed)
  57+13+0 (1/0/0/0) _top_
  79+1+0 (1/0/0/0) Section: Abstract
  1385+1+235 (1/2/5/0) Section: Letter
  56+1+0 (1/0/0/0) Section: Acknowledgements
  0+2+0 (1/0/0/0) Section: Word count
~~~

## References

[1] F L Mackie, K Hemming, S Allen, R K Morris, and M D Kilby. The accuracy of cell-free fetal dna-based non-invasive prenatal testing in singleton pregnancies: a systematic review and bivariate meta-analysis. BJOG, 124(1):32–46, Jan 2017.

[2] Jinglan Zhang, Jianli Li, Jennifer B Saucier, Yanming Feng, Yanjun Jiang, Jefferson Sinson, Anne K McCombs, Eric S Schmitt, Sandra Peacock, Stella Chen, Hongzheng Dai, Xiaoyan Ge, Guoli Wang, Chad A Shaw, Hui Mei, Amy Breman, Fan Xia, Yaping Yang, Anne Purgason, Alan Pourpak, Zhao Chen, Xia Wang, Yue Wang, Shashikant Kulkarni, Kwong Wai Choy, Ronald J Wapner, Ignatia B Van den Veyver, Arthur Beaudet, Sheetal Parmar, Lee-Jun Wong, and Christine M Eng. Non-invasive prenatal sequencing for multiple mendelian monogenic disorders using circulating cell-free fetal dna. Nat Med, Jan 2019.

[3] David S Tsao, Sukrit Silas, Brian P Landry, Nelda P Itzep, Amy B Nguyen, Samuel Greenberg, Celeste K Kanne, Vivien A Sheehan, Rani Sharma, Rahul Shukla, Prem N Arora, and Oguzhan Atay. A novel high-throughput molecular counting method with single base-pair resolution enables accurate single-gene nipt. Sci Rep, 9(1):14382, Oct 2019.

[4] L J Salomon, A Sotiriadis, C B Wulff, A Odibo, and R Akolekar. Risk of miscarriage following amniocentesis or chorionic villus sampling: systematic review of literature and updated meta-analysis. Ultrasound Obstet Gynecol, 54(4):442–451, Oct 2019.

[5] Jacob O Kitzman, Matthew W Snyder, Mario Ventura, Alexandra P Lewis, Ruolan Qiu, Lavone E Simmons, Hilary S Gammill, Craig E Rubens, Donna A Santillan, Jeffrey C Murray, Holly K Tabor, Michael J Bamshad, Evan E Eichler, and Jay Shendure. Noninvasive whole-genome sequencing of a human fetus. Sci Transl Med, 4(137):137ra76, Jun 2012.

[6] Matthew W Snyder, LaVone E Simmons, Jacob O Kitzman, Donna A Santillan, Mark K Santillan, Hilary S Gammill, and Jay Shendure. Noninvasive fetal genome sequencing: a primer. Prenat Diagn, 33(6):547–554, Jun 2013.

[7] Sarah L Kinnings, Jennifer A Geis, Eyad Almasri, Huiquan Wang, Xiaojun Guan, Ron M McCullough, Allan T Bombard, Juan-Sebastian Saldivar, Paul Oeth, and Cosmin Deciu. Factors affecting levels of circulating cell-free fetal dna in maternal plasma and their implications for noninvasive prenatal testing. Prenat Diagn, 35(8):816–822, Aug 2015.

[8] K C Allen Chan, Peiyong Jiang, Kun Sun, Yvonne K Y Cheng, Yu K Tong, Suk Hang Cheng, Ada I C Wong, Irena Hudecova, Tak Y Leung, Rossa W K Chiu, and Yuk Ming Dennis Lo. Second generation noninvasive fetal genome analysis reveals de novo mutations, single-base parental inheritance, and preferred dna ends. Proc Natl Acad Sci USA, 113(50):E8159–E8168, Dec 2016.

